# Predicting COVID19 Critical Care Beds - The London North-West University Healthcare Trust Experience

**DOI:** 10.1101/2020.11.20.20235226

**Authors:** J.A Naude, M. Taniparthi, K. Adams, J. Biggin-Lamming, R. Elias, P. Milner, I. Mohammed, M. Towers, M. Kuper

## Abstract

Our trust has an urgent need to make short-term (3-4 days in advance) informed operational decisions which take into account best-practice treatment regimens and known clinical features of COVID19 inpatients.

We believe that any model which is relied upon for operational decision making should have clinically identifiable parameters. Our model’s parameters take into account the conversion rates from acute wards into wards equipped with Non-Invasive Ventilation (NIV) and Mechanical Ventilation (MV), the typical time that these conversions take place and, the historical non-COVID usage of NIV and MV beds.

We have observed that this clinical performance is mathematically identical to a series of linear delays on the time varying inpatient level. High frequency inpatient data, sampled ∼4 hourly, has allowed our hospital trust to predict total critical care usage up to 4 days in advance without making any assumptions on upcoming inpatients. It is based entirely upon current bed occupancy levels and measured clinical pathways.

Through back-testing over the recent 4 months, the bounds of this model include 93.8% of all 4 day inpatient sequences. The average next-day error is 0.8 (95% CI: 0.44, 1.15) and so the system tends to over-predict the next day critical care inpatients by approximately 1 bed.

Potential extensions to the basic model include adjustments for seasonality, case mix, probabilistic marginalisation and known discharges.

## Method

The line of reasoning involves bed occupancy as a key signal, denoted as *x*(*t*).

We have reliably measured a scaling and time shift of COVID bed occupancy with respect to NIV and MV needs. We note ∼ 18% of COVID inpatients require Non-Invasive Ventilation (NIV) and ∼ 6% require Mechanical Ventilation (MV). As an ensemble average, these inpatients require these treatments at 4 and 6 days respectively.

Hence, if COVID bed occupancy levels are currently known, the probable critical care inpatients can be reasonably estimated as 24% of these. We would also expect any changes in COVID inpatient levels to be reflected within 4 - 6 days’ time.

Mathematically, the baseline estimate for total critical care, denoted *ŷ*(*t*) is given by

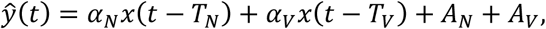

where

*α*_*N*_ is the fraction of COVID positive bed occupancy that convert to NIV,

*α*_*V*_ is the fraction of COVID positive bed occupancy that convert to MV,

*T*_*N*_ is the measured delay in conversion to NIV,

*T*_*V*_ is the measured delay in conversion to MV,

*A*_*N*_ is the historical average non-COVID NIV bed occupancy,

and, *A*_*V*_ is is the historical average non-COVID MV bed occupancy.

The prediction interval includes upper and lower bounds above and below this baseline by incorporating twice the average deviation of the historical non-COVID critical care bed occupancy.

In order to implement this solution, it was necessary to have high quality and frequent updates to the COVID inpatient count and the trust’s electronic patient record system was configured to have database views that were dedicated to this task.

### Fractional Inpatient Levels

The fractional parameters were fit using a ratio of running total of COVID positive NIV inpatients to running total of COVID positive inpatients over time since 1 September 2020 i.e.

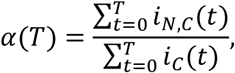

where *i*_*N,C*_(*t*) is the COVID positive NIV inpatient level at sample instant *t* and *i*_*C*_(*t*) is the total COVID positive inpatient level at sample instant *t*. For comparison, the parameters were also fit with a time average of the instantaneous ratio i.e.

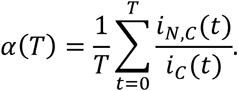

This second method is subject to large variation due to the low levels at the start of the second wave and these are depicted in Figure 1.

**Figure 1:**
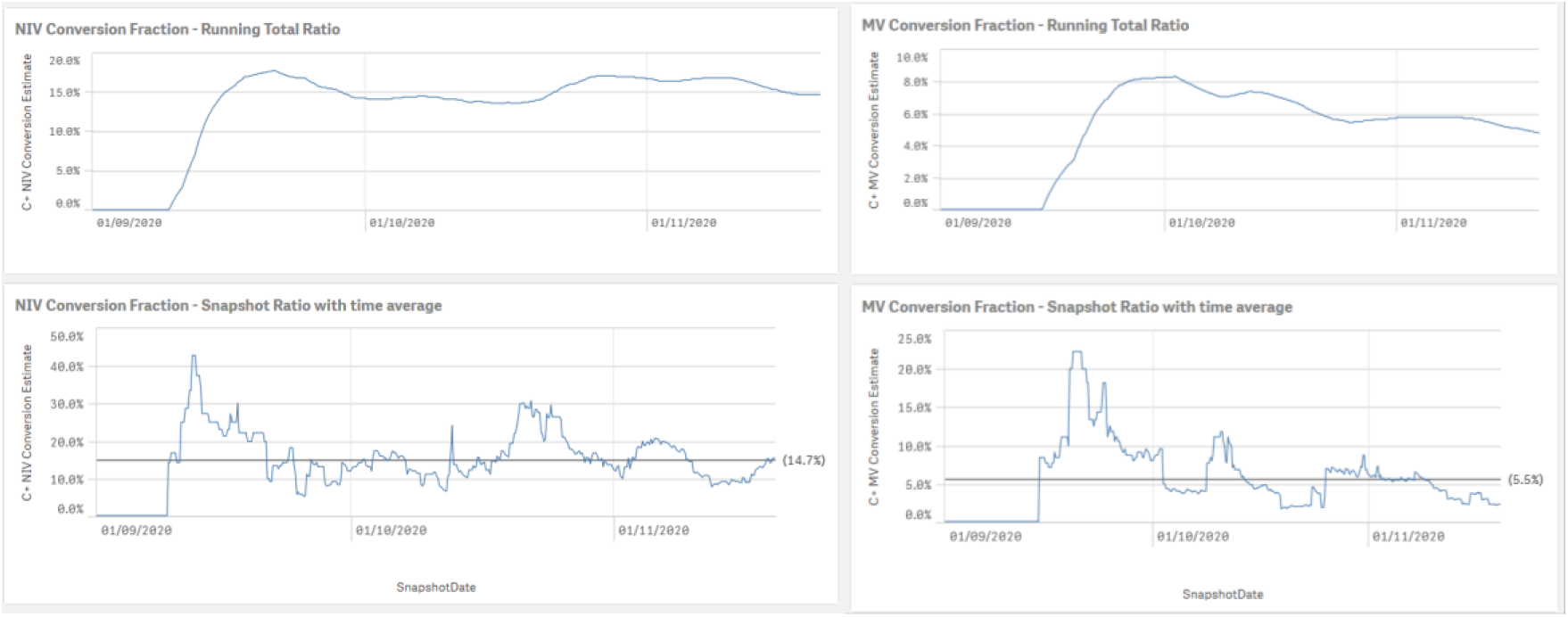
Fractional Inpatient Conversion.

### Time Delay to Conversion

The time parameters were fit by using the patient ensemble average of minimum time between admission and being present within the NIV or MV category respectively. There will be some uncertainty as to the exact time to conversion due to operational data capturing and the 4 hourly database snapshot. The ensemble included 70 NIV and 15 MV COVID positive conversions since 1 Sept 2020.

The results are *T*_*N*_ = 3.41(95% CI: 2.34, 4.47) and *T*_*V*_ = 6.42 (95% CI: 4.77, 8.06) respectively. Given the need for integers due to the data samples, 4 and 6 were chosen respectively as good first approximations.

## Results

Database snapshots from the present 4 hourly interval extending backwards to late June 2020 were immediately available within the trust. The tool correctly predicted the critical care needs during a second wave COVID surge in inpatient levels in mid-October 2020. This is depicted in Figure 2, where the left hand side is the actual critical care inpatient level and the right hand side are the predictions for 4 days in advance. Note that *every value* on the right hand side of Figure 2 is a prediction from the linear delay model as a function of total COVID positive inpatient levels.

**Figure 2:**
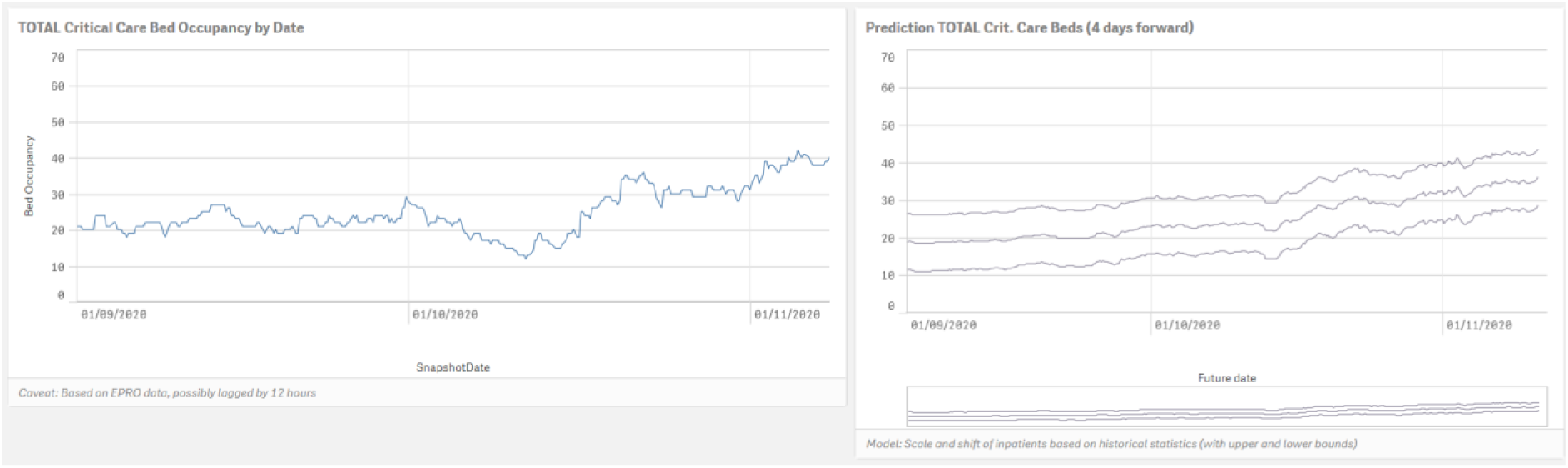
Screenshot of prediction tool [Sept to mid Nov. 2020].

Most importantly, during the middle of October where actual critical usage was decreasing, the model correctly anticipates a surge to a new level of approximately 30 critical care beds before this event materialises.

The above results utilise the clinically derived parameters and the bounds of the prediction included 93.8% of all 4 day actual inpatient levels over the 4 months of historical data. The average next-day error is 0.8 (95% CI: 0.44, 1.15) patients over the full time frame.

As a test, the model structure was frozen and the parameters were fit with a generalised reduced gradient algorithm, incorporating integer constraints on the delays and the requirement that the MV fraction be below the NIV fraction. A multi-start run with 100 initial conditions was employed and a Bayesian stopping criterion was used. The resulting parameters from this exercise were near identical to the clinically derived parameters and gave no additional performance with regards to the fraction of actuals contained within the prediction bounds.

## Discussion

Measuring *x*(*t*) automatically includes admissions, discharges and length of stay. For example, if bed occupancy decreases, then either there are fewer admissions or more discharges or length of stay is reduced or some combination of all of these effects.

This line of thinking is subtle. The underlying clinical situation may be rapidly evolving, though the bed occupancy could remain constant. This simple relationship is obfuscated when looking at admitted cohorts, length of stay and discharges and how these change over time. The bed occupancy as a function of time appears to be a sufficient statistic. A theoretical exposition is intended to follow this rapid communication in a follow-up paper.

It is noteworthy that brute force numerical optimisation of the parameters within the model resulted in much the same values as those clinically derived.

The results are encouraging and the system is currently being used to assist with operational decision making within the trust to safe guard our critical care patients and ensure adequate supply for the variation in expected short term needs.

### Refinements

The average levels for NIV and MV, *A*_*N*_ and *A*_*V*_ depend on historical conversion ratios of non-COVID inpatients. Each of these average levels can be further sub-divided to incorporate the inpatient case-mix for other diseases. This would extend the methodology to multiple diseases and if accurate records are timeously available on these inpatient levels, critical care utilisation may be more generally predictable.

Stated differently, the current case-mix labelling is currently for COVID and non-COVID inpatients only. There is no reason that the non-COVID inpatients may be further sub-divided into their various case types and, provided there is a typical delay before needing critical care, the same anticipatory action may be employed to predict a changing case mix on the critical care facilities available.

The system currently employs a single fraction and delay time for each critical care component. This may be extended by, in effect, probabilistically marginalising over the various fractions and timings of sub-cohorts of the patient ensemble. For example, if 15% overall inpatients convert to NIV at around 4 days; it is likely that some smaller fraction will convert in sooner than 4 days, and another fraction will convert later than 4 days so that overall these parameters are valid. There is ample evidence that this state of affairs matches reality as reported by clinical staff who are presently engaged with treatment.

As another possible refinement on other case mixes; the seasonality of the historical critical care usage may be incorporated if there is insufficient delay between inpatient levels rising and critical care levels rising.

Transfers (clinical and non-clinical) from critical care act as a disturbing input to the steady state levels predicted with this model. The model was developed at a time of few transfers out from critical care due to capacity reasons so no specific allowance has been made for this. As a refinement for the short-term predicted level, these transfers may be subtracted from the steady state predictions to enhance next-day accuracy.

It is hoped that this framework stimulates healthcare practitioners, operational managers and other members within our healthcare community to extend the current practice regarding anticipated need. We believe that only by marrying clinical insight with robust and clinically transparent mathematics will the results be trusted and the full promise of our current data be realised.

### Limitations

This methodology can be reasoned to be ‘deductive’ in the sense that the prediction time horizon is intimately dependent on observed delays in the clinical evolution of inpatient numbers. If the time shift was not at all present, then future needs would depend on future inpatient numbers (which would be probabilistic). Currently, the system is configured to use up to the present patient numbers and hence the prediction time horizon is dependent on observed delays.

